# Optimal processing of tongue swab samples for Mycobacterium tuberculosis detection by the Xpert MTB/RIF Ultra assay

**DOI:** 10.1101/2024.06.20.24309244

**Authors:** Gayatri Shankar Chilambi, Robert Reiss, Naranjargal Daivaa, Padmapriya Banada, Margaretha De Vos, Adam Penn-Nicholson, David Alland

## Abstract

Tongue swabs represent a potential alternative to sputum as a sample type for detecting pulmonary tuberculosis (TB) using molecular diagnostic tests. The methods used to process tongue swabs for testing in the WHO-recommended Xpert MTB/RIF Ultra (Xpert Ultra) assay vary greatly. We aimed to identify the optimal method for processing diagnostic tongue swabs for subsequent testing by Xpert Ultra. We compared four methods for treating dry tongue swabs with Xpert Ultra sample reagent (SR) mixed with various concentrations of Tris-EDTA-Tween (TET), to treatment with SR alone or to a commonly used SR-free heat-inactivation protocol. In each condition, swabs obtained from volunteers without TB were placed into test buffer spiked with known amounts of *Mycobacterium tuberculosis* (*Mtb*) strain H37Rv-mc^2^6230. Swabs processed with 1:1 diluted SR buffer had the lowest *Mtb* limit of detection (LOD) at 22.7 CFU/700 µl (95% CI 14.2-31.2), followed by 2:1 diluted SR buffer at 30.3 CFU/700 µl (95% CI 19.9-40.7), neat SR at 30.9 CFU/700 µl (95% CI 21.5-40.3) and SR prefilled in the Xpert Ultra at 57.1 CFU/700 µl (95% CI 42.4-71.7). Swabs processed using the heat-based protocol had the highest LOD (77.6 CFU/700 µl; 95% CI 51.2–104.0). Similar findings were observed for LOD of RIF-susceptibility. Assay sensitivity using the 2:1 diluted SR buffer did not vary considerably in the presence of sputum matrix or phosphate buffer saline. Further studies are needed to assess the performance of this processing protocol in a clinical setting.

**Importance:** Xpert MTB/RIF Ultra (Xpert Ultra) is approved by the World Health Organization for the diagnosis of tuberculosis (TB). This test is typically performed using sputum specimens obtained from people with presumptive TB. In order to inactivate *Mtb* and aid liquefaction, sputum must be mixed with Xpert SR prior to transfer into the Xpert Ultra. However, some people under evaluation for TB are unable to produce sputum. Alternative sample types for TB diagnosis would therefore be of value. Oral-swabs, including tongue-swabs have shown promise, but there are technical challenges associated with sample processing. In this study, several new tongue swab processing conditions were evaluated, utilizing SR, either neat or diluted in buffer. The ability of Xpert Ultra to detect TB was improved under these conditions compared with the previously published heat-processing method (1–3), processing steps were simplified, and technical challenges were overcome.

## Introduction

Tuberculosis (TB) remains a major public health threat. Globally, 7.5 million people were newly diagnosed with TB in 2022 (4); even though an estimated 10.6 million actually developed TB during that year, leaving 3.1 million people undiagnosed or underreported (4). Delays in diagnosis contribute to low TB treatment rates and consequently to increased mortality and disease transmission (4, 5).

Most existing TB diagnostics rely on sputum specimens for *Mycobacterium tuberculosis* (*Mtb*) complex detection (5). These include sputum-smear microscopy, culture-based tests, or molecular WHO-recommended diagnostic (mWRD) tests such as Xpert MTB/RIF and Xpert MTB/RIF Ultra (Xpert Ultra; Cepheid, Inc., Sunnyvale, CA); the Truenat assays MTB, MTB Plus and MTB RIF-Dx (Molbio Diagnostics Ltd, Verna, Goa, India) and moderate complexity assays such as Abbott RealTi*m*e MTB assays (Abbott Laboratories, Abbott Park, USA), Roche Cobas MTB (Hoffmann-La Roche, Basel, Switzerland), Fluorotype MTBDR (Bruker/Hain Lifescience, Nehren, Germany), BD MAX MDR-TB (Becton, Dickinson and Company, Franklin Lakes, USA) and the TB-LAMP assay (Eiken Chemical, Tokyo, Japan) (5). However, TB diagnosis with sputum-based technologies is associated with challenges, including the inability of some patients to produce sputum, such as younger children or people living with HIV (6–10). Several non-sputum-based diagnostics are under investigation to address these needs, utilizing specimen types such as blood, stool, urine, and breath aerosols.

Considerable effort has been undertaken studying the potential of tongue swabs as an alternative non-invasive oral sample type for pulmonary tuberculosis (1, 2). Previous studies with tongue swabs have reported 52-91% sensitivity in adults, with considerable variation according to sample processing methods, sampling site, swab type, storage conditions, and analysis approaches (2). These studies evaluated the ability of automated TB detection platforms such as Xpert Ultra (1, 3, 11–13) and MTB Ultima (Molbio Diagnostics Ltd, Verna, Goa, India) (3), or in-house PCR tests (14), to detect *Mtb* in tongue swabs either spiked with *Mtb* bacilli or confirmed TB positive by a reference standard.

Xpert Ultra is known to have a low limit of detection (LOD), and has optimal analytical sensitivity to detect *Mtb* in non-sputum specimens such as cerebrospinal fluid (CSF), lymph, and tissue biopsies using processing protocols based on the Cepheid proprietary Xpert sample reagent (SR) (5, 15–18). Few studies have assessed the TB detection when tongue swabs were processed with Xpert Sample Reagent (SR) based on the existing sputum based Xpert MTB/RIF Ultra (Ultra) protocol (1, 11). Andama *et al* showed analytically with spiked tongue swabs that a single tongue swab treated with SR yielded a higher LOD in comparison to processing double swab with SR or single swab with a heat-based processing approach. However, isolated reports of over-pressurization errors (19) are observed with Xpert Ultra testing of tongue swab samples based on current heat-based sample processing protocols. Developing an SR-based tongue swab processing protocol optimized for Xpert Ultra testing, that does not result in over-pressure errors, and enables improved assay sensitivity would be helpful.

The objective of this study was to evaluate the impact of four modifications of tongue swab sample processing conditions with different dilutions of Xpert SR on the performance of Xpert Ultra for detection of *Mtb* and rifampicin (RIF) resistance, when compared with the existing heat-based inactivation protocol (1) for the Xpert Ultra assay. The impact of tongue biomass, sputum matrix swab, and buffer on performance using one selected sample processing condition was also evaluated.

## Materials and methods

### Study participants and swab sample collection

Healthy volunteers (aged > 18 years) working at the International Centre for Public Health at Rutgers University in New Jersey, USA, were recruited by flyer, mobile or email notifications for this study. Participants were asked not to eat or drink, use tobacco or oral hygiene products, or expectorate sputum up to 30 minutes before collecting their tongue swabs. Participants were provided with sterile, single-use, and individually packaged Copan FLOQSwabs (520CS01) to self-collect, rolling the swab head along the breadth of their mid-tongue or back half of the dorsum of the tongue without touching their teeth or throat, within 10 seconds. After using a single FLOQSwab, the swab head was snapped at the 30 mm breakpoint into a 2 ml sterile and empty screw cap collection tube. The closed tubes were placed inside a cooler box in a leak-proof biohazard bag. All the swabs were stored at 2-8°C for less than a week before testing.

In order to evaluate the impact of sputum matrix swabs on Xpert Ultra performance, sputum from leftover clinical specimens collected from the University Hospital Microbiology Lab (University Hospital, Newark, NJ, USA) was used.

This study was approved by the Rutgers Institutional Review Board (IRB protocol # Pro2021002454). The use of sputum from leftover clinical specimens was approved by the Rutgers Institutional Review Board, IRB protocol numbers 020160000657 and 0120090104.

### Preparation and quantitation of concentrated viable Mtb cell stock

Titered stocks of an attenuated version of the *Mtb* H37Rv (mc^2^6230) laboratory strain were prepared and quantified as previously described (17). The *Mtb* cells were inoculated in a ratio of 1:100 in 10 mL of 7H9 broth supplemented with 10% Middlebrook OADC Growth supplement, 0.05% Tween 80 (Sigma Aldrich, St Louis, MO), and 24 µg/ml of calcium pantheonate (Sigma Aldrich, St Louis, MO). *Mtb* grown to an optical density at 600 nm (OD600) of 0.6–0.8 was sub-cultured twice before dilutions were prepared to quantify colony-forming units (CFU); the culture was then divided into 500 µl aliquots and stored at −80°C until used. The 10^-5^, 10^-6,^ and 10^-7^ dilutions were plated in triplicate on 7H10 agar plates supplemented with 10% Middlebrook OADC Growth supplement and 24 µg/mL calcium pantheonate. Plates were incubated for two to three weeks at 37°C, and the CFU/ml of the culture stock was estimated from plates with colony counts ranging between 300 and 10.

### Cell preparation and dilution for spiking

Cell dilutions were prepared as previously described (17). The quantified stocks of *Mtb* were removed from −80°C storage and thawed on ice. Thereafter, 500 µl of supplemented 7H9 media was added to a 500 µl aliquot of the quantified *Mtb* stock, which was mixed by vortexing for 30 seconds at medium speed and placed on ice for 6 minutes to allow aerosolized particles to settle. The mixture was then sonicated, and serial dilutions between 10 and 10^7^ CFU/mL were made with supplemented 7H9. The stock suspensions were stored at 4°C and sonicated once for 30 seconds on the subsequent day. Stock suspension used on additional days (days 3–7) was vortexed at medium speed for 2 minutes and restricted to a maximum of one such vortex every 24 hours.

### Spiking and sample processing conditions

The tongue swabs of human volunteers were placed in 700 µl of buffer either spiked or unspiked with known *Mtb* CFUs and thereafter processed using five different sample processing conditions, referred to as conditions 1–5 (**Fig.1**). Each of these protocols tested included either a Tris-EDTA-Tween buffer (TET, 50 mM Tris, 0.1 mM EDTA, 0.1% Tween20, pH 8.4) or a combination of TET buffer and SR as the sample processing reagents. Each sample processing condition was tested simultaneously, with final concentrations of *Mtb* of 0, 7, 14, 28 and 56 CFU/700 µl. Conditions 1 and 5 were tested at two additional concentrations of 112 and 336 CFU/700 µl.

**FIG 1.**
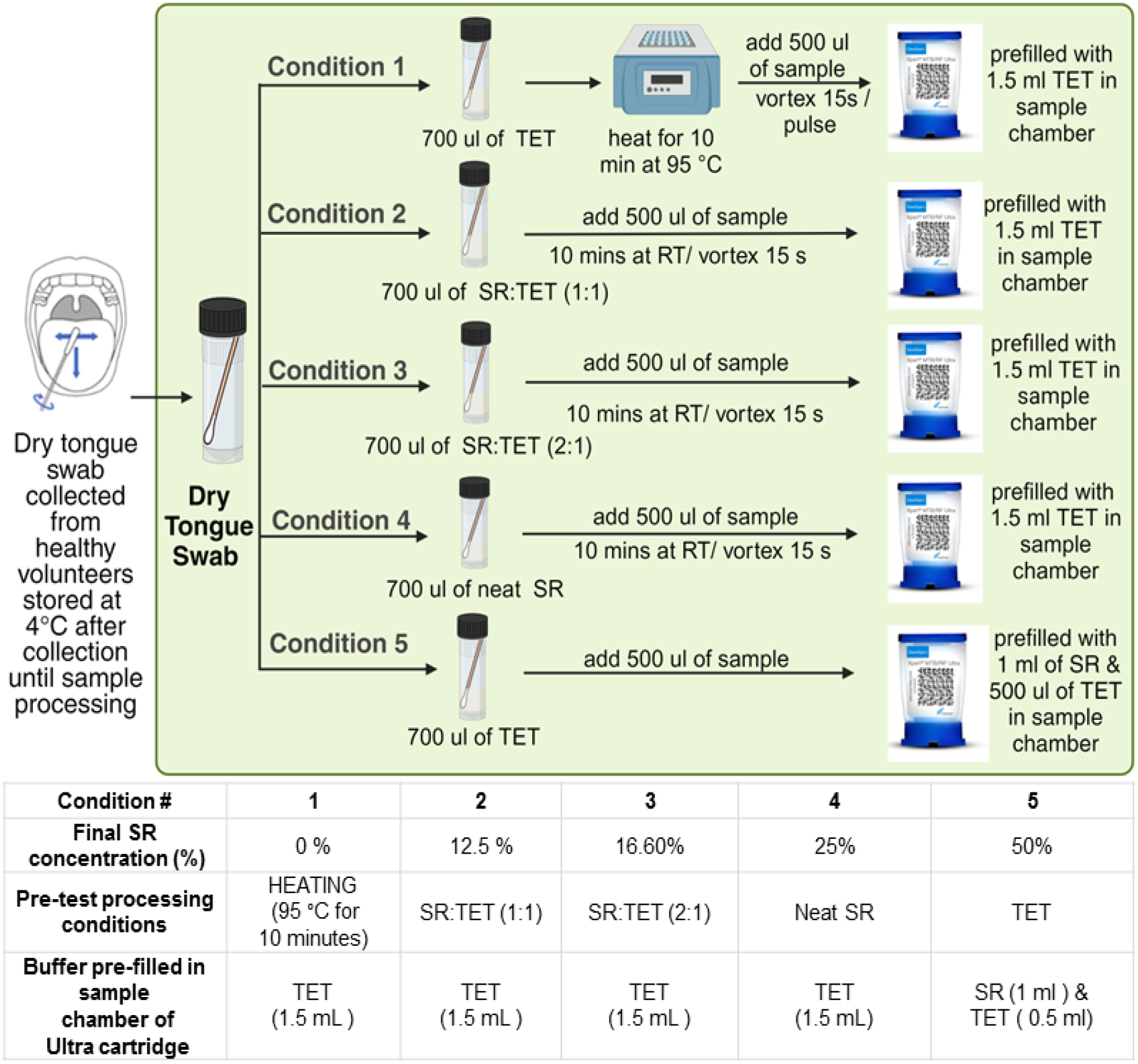
Sample processing workflow showing how tongue swabs (spiked or unspiked) were processed before loading into the Xpert MTB/RIF Ultra cartridge. RT, room temperature; SR, Xpert sample reagent; TET, Tris-EDTA-Tween buffer. Created with BioRender.com.

Condition 1 was the existing heat-based inactivation protocol for the Xpert Ultra assay and was used as a comparator in this study. In condition 1, 700 µl of Tris-EDTA-Tween buffer spiked with *Mtb* was transferred by a pipette to the tube containing the swab head and heated for 10 minutes at 95°C. Samples were cooled to room temperature before vortexing for 15 seconds and pulse centrifugation. Subsequently, 500 µl of the supernatant was transferred into the Xpert Ultra cartridge sample chamber prefilled with 1.5 mL of TET buffer. In sample processing conditions 2, 3, and 4, 700 µl of SR and TET buffer were prepared in a 1:1 or 2:1 ratio, or neat SR, respectively, with the 2:1 ratio representing the SR:sample proportions used for treating sputum samples with Ultra. These 700 µl buffer aliquots were then spiked with *Mtb* and transferred to the tube containing the swab head, vortexed briefly and incubated at room temperature. After 10 minutes of incubation, 500 µl of the sample was transferred to the Xpert Ultra cartridge sample chamber prefilled with 1.5 mL of TET buffer. Finally, in condition 5, 700 µl of TET buffer spiked with *Mtb* was added to the tube containing the swab, and 500 µl of the sample from the tube was loaded into the Xpert Ultra cartridge prefilled with 1 ml of SR and 500 µl of TET buffer. Condition 5 was designed to test assay performance where tongue swab heads that were exposed briefly to higher final concentrations of SR than conditions 2-4.

### Xpert Ultra performance assessment

Xpert Ultra performance under each sample processing condition was assessed according to the *Mtb* dilution until *Mtb* positivity or RIF resistance could be detected in 100% of samples. Any test that resulted in an invalid or error output from the GeneXpert instrument was excluded from the primary analysis but recorded for error reporting. The LOD under each condition was also calculated, defined as the lowest concentration of the analyte (IS*6110*/IS*1081* for *Mtb* positivity and *rpoB* for RIF susceptibility) detected with 95% certainty (17). Mean IS*6110*/IS*1081* and *rpoB* cycle threshold (Ct) values were measured for each condition.

One sample processing condition was then selected based on associated LOD and biosafety considerations for further analyses, which included the impact of tongue biomass, sputum matrix swab, and use of phosphate buffer saline (PBS) instead of Tris-EDTA-Tween, on Xpert Ultra performance under the selected condition. In-cartridge pressure values of Xpert Ultra tests performed with tongue and sputum matrix swabs were noted.

### Statistical analysis

All statistical analyses were performed using R Statistical Software version 4.3.1 (R Foundation for Statistical Computing, Vienna, Austria) or GraphPad Prism Version 9.5.1 (733).

Probit regression was used to model the detection rate as a function of concentration and LODs were calculated using inverse estimation. LOD estimates for two conditions were considered to be significantly different if there was no overlap between the Wald 95% confidence intervals for each condition.

Two-way ANOVA was used to examine the differences in Ct values between concentrations under multiple sample processing conditions. The Ct values of condition 1 and 5 were compared using independent sample t-tests at concentrations greater than 56 CFU/700ul. Only Ct values of samples positive for *Mtb* (in case of IS*6110*/IS*1081* Ct) (Fig. 3a) and RIF susceptibility (in case of *rpoB* Ct) (Fig. 3b) were considered for statistical analysis. Post-hoc pairwise comparisons were conducted using Tukey’s multiple comparisons test.

For conditions 1 and 3, the medians of in-cartridge pressure values were compared using Mood’s median test, and the means of the Ct values were compared using independent sample t-tests. Homogeneity of variance for each condition was tested using Levene’s test. Detection rates under differing conditions were compared using Fisher’s exact test.

## Results

### Sample processing conditions assessed by limit of detection

We determined the limit of detection (LOD) of samples processed using the five sample processing conditions and then tested with Xpert Ultra assay to estimate the influence of the different tongue swab sample processing conditions on the assay performance. Xpert Ultra assay results provide information on the presence of *Mtb,* and the presence of RIF susceptibility or resistance based on their respective molecular targets IS*6110*/IS*1081* and *rpoB*. *Mtb* detection LOD is usually more sensitive as observed in cases where IS*6110*/IS*1081* is detected but *rpoB* remains undetected in the sample (17). For *Mtb* detection, the LOD (**Fig. 2a**) was lowest for swabs processed by condition 2 at 22.7 CFU/700 µl (95% CI 14.2-31.2), followed by conditions 3 and 4 at 30.3 CFU/700 µl (95% CI 19.9-40.7) and 30.9 CFU/700 µl (95% CI 21.5-40.3), respectively.

**FIG 2.**
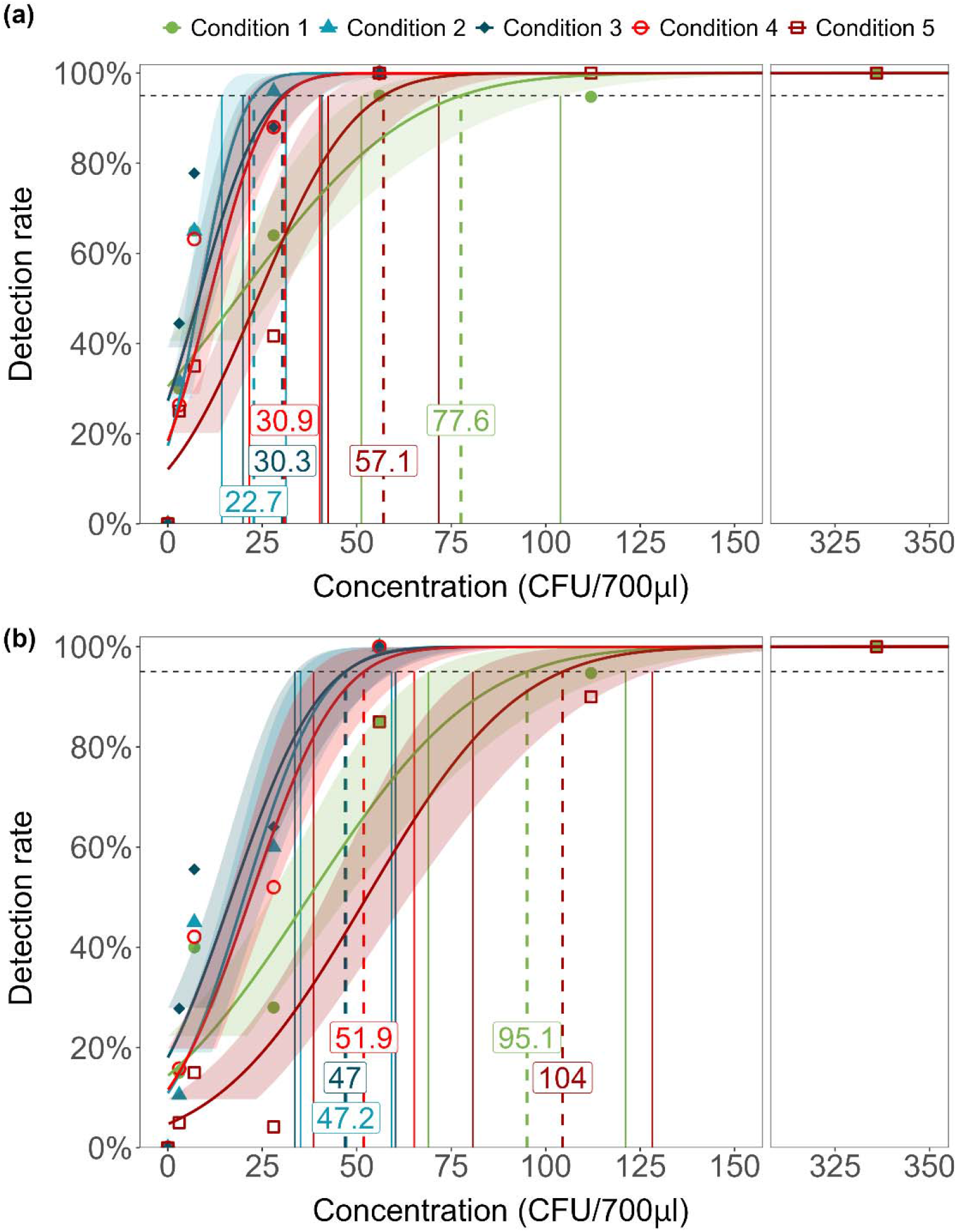
Limit of detection (LOD) of (a) *Mtb* positivity and (b) RIF susceptibility for tongue swabs under sample processing conditions 1–5. Dashed lines indicate concentration at the 95% limit of detection, and solid lines indicate the upper and lower bounds of Wald 95% confidence interval. The numbers in the boxes are labels for concentration at 95% LOD for each condition. CFU, colony forming unit.

The LOD of *Mtb* with swabs processed by condition 5 was 57.1 CFU/700 µl (95% CI 42.4-71.7). Swabs processed by condition 1 had the highest LOD of 77.6 CFU/700 µl (95% CI 51.2-104.0).

For detection of RIF susceptibility or resistance, the LOD was based only on whether the output of the Xpert Ultra assay reported RIF as susceptible since no RIF resistant isolates were tested. Assays that were reported as trace or RIF indeterminant were both considered negative in the LOD calculations. (**Fig. 2b**), the LODs of condition 2 (47.2 CFU/700 µl [95% CI 35.1-59.2]), condition 3 (47.0 CFU/700 µl [95% CI, 33.7-60.2]) and condition 4 (51.9 CFU/700 µl [95% CI 38.5-65.2]) were all comparable. A higher LOD of RIF susceptibility was observed in swabs processed by condition 1 (95.1 CFU/700 µl [95% CI 68.9-121.2]) and condition 5 (104.5 CFU/700 µl [95% CI 80.7-128.3]) (**Fig. 2b**). No false positive results for *Mtb* detection or RIF resistance were observed in any of the samples tested.

### Performance of Xpert Ultra by sample processing condition based on cycle threshold values

The IS*6110*/IS*1081* (for Mtb detection) and *rpoB* (for RIF susceptibility detection) Ct values resulting from our LOD studies of tongue swab samples processed by condition 1–5, were plotted based on the numbers of *Mtb* CFU spiked into each sample condition (**Fig. 3**). A significant difference was observed in the mean IS*6110*/IS*1081* Ct values of condition 1 and 5 for samples tested at 336 CFU/700 µl (p<0.0001) and 112 CFU/700 µl (p =0.0406). IS*6110*/IS*1081* Ct values of samples tested at 56 CFU/700 µl with condition 5 were significantly different from condition 1 (p = 0.0409), condition 2 (p< 0.0001), condition 3 (p=0.0007) and condition 4 (p=0.0021). A significant difference was observed in the mean *rpoB* Ct values in condition 1 and 5 for samples tested at 336 CFU/700 µl (p = 0.0024) and 112 CFU/700 µl (p = 0.0036). At 56 CFU/700 µl, a significant difference was observed between the mean *rpoB* Ct values of samples processed by condition 5 versus condition 1 (p=0.0007), condition 2 (p<0.0001), condition 3 (p<0.0001) and condition 4 (p=0.0002). However, there was no significant difference between *rpoB* Ct values in samples tested at concentrations below 56 CFU/700 µl for any sample processing method.

**FIG 3.**
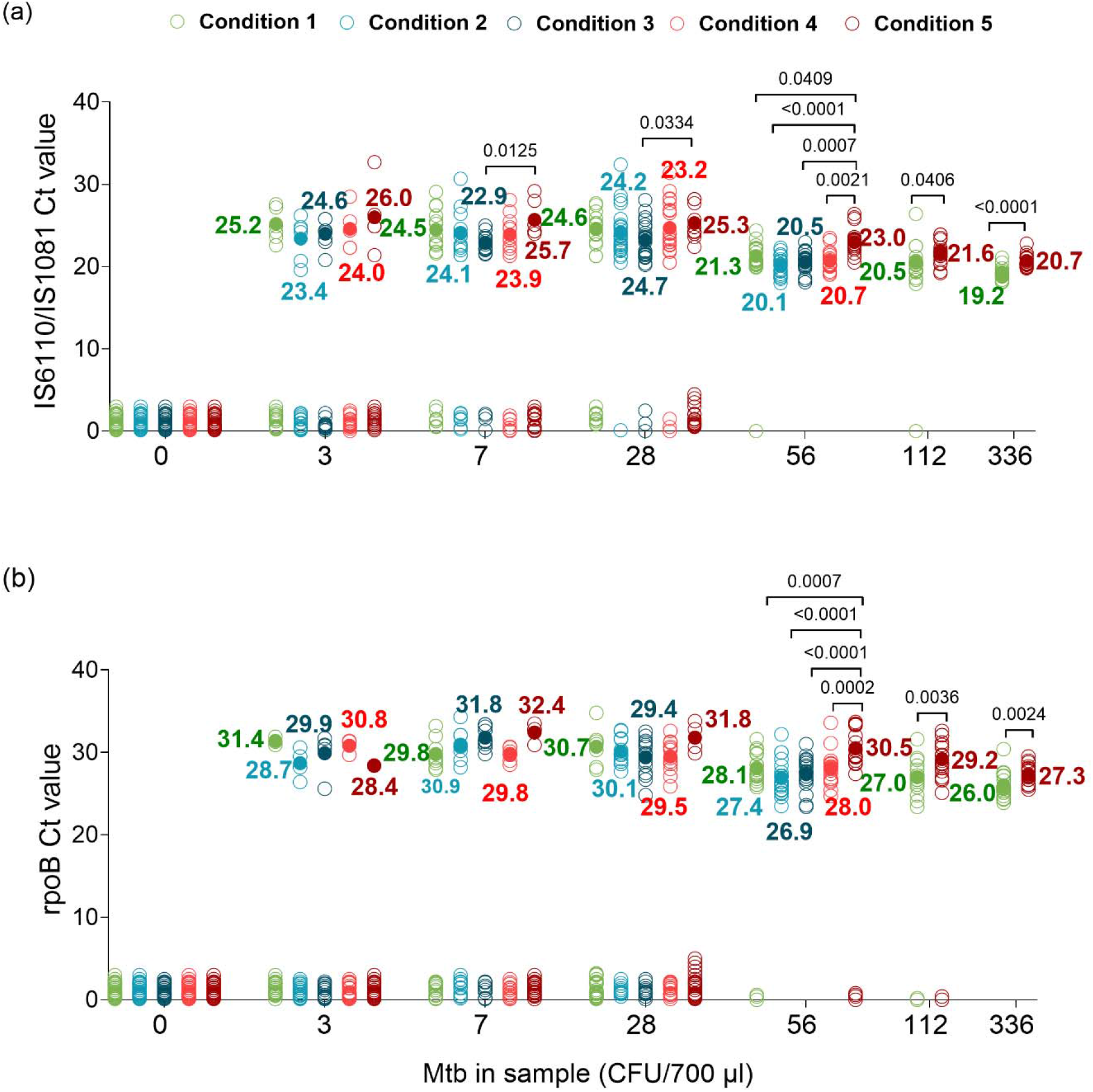
Ct values for (a) IS*6110*/IS*1081* (b) *rpoB* for tongue swabs under sample processing conditions 1–5. Mean Ct values are indicated by filled circles. CFU, colony forming unit; Ct, cycle threshold; *Mtb*, *Mycobacterium tuberculosis*.

### Selecting the optimal swab treatment condition

Based on the combined LOD and Ct value findings (**Table 1**, **Fig 2**, **Fig 3**) of both the *Mtb* and rifampin resistance components of the assay, conditions 2, 3 and 4 were not statistically different. However, we considered the fact that condition 3 had modestly higher CFU detection rates in samples spiked with lower numbers of CFU (3, and 7 CFU for *Mtb* detection and 3, 7, and 28 CFU for rifampin susceptibility detection, **Table 1**), along with prior evidence that 2:1 dilutions of SR:sample effectively decontaminate samples (20). These considerations led us to select condition 3 (SR: Tris-EDTA-Tween buffer 2:1) as the optimal tongue swab processing method for further analytical studies. Condition 1 (existing heat-based protocol) continued to be used as a comparator.

**TABLE 1.**
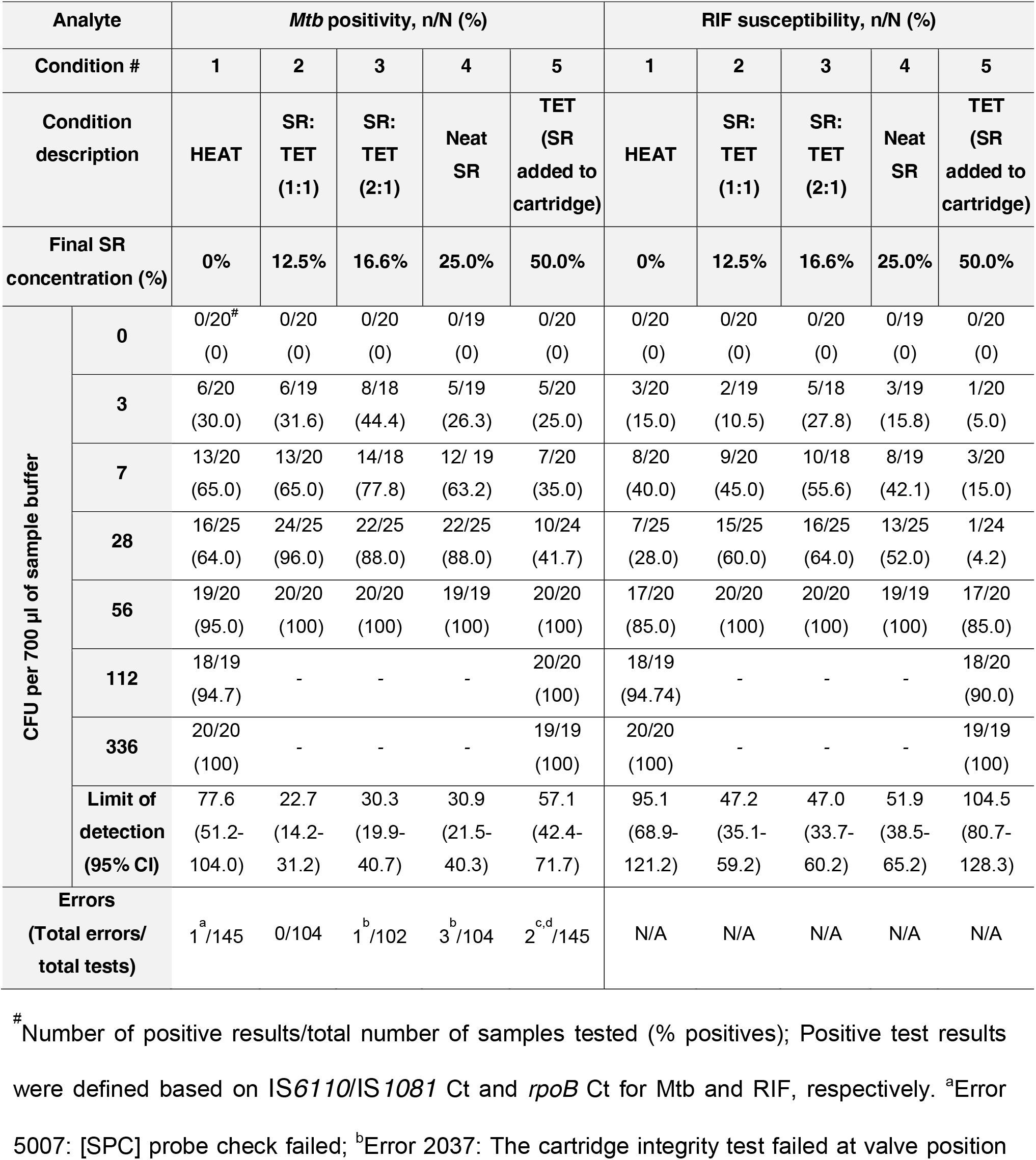
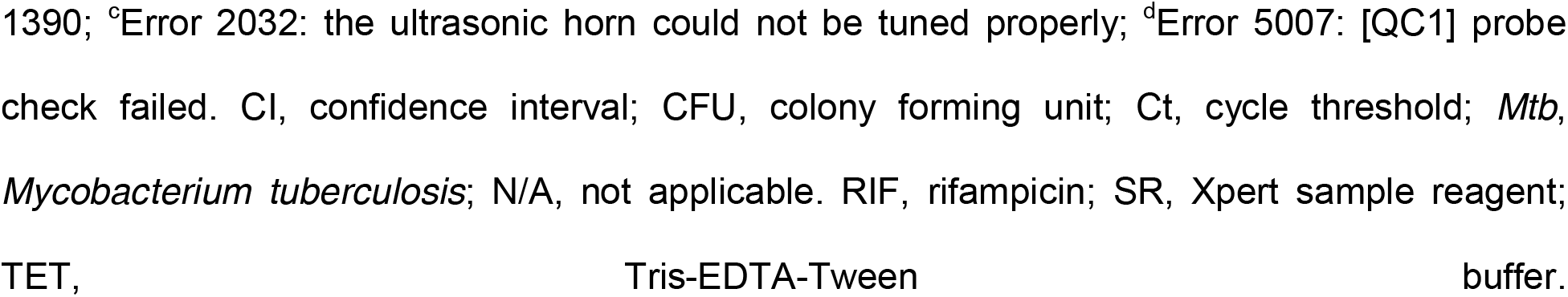
Comparative assessment of Xpert Ultra assay testing of *Mtb* spiked into buffer with tongue swabs by sample processing condition.

| Analyte |  | *Mtb* positivity, n/N (%) |  |  |  |  | RIF susceptibility, n/N (%) |  |  |  |  |
| --- | --- | --- | --- | --- | --- | --- | --- | --- | --- | --- | --- |
| Condition # |  | 1 | 2 | 3 | 4 | 5 | 1 | 2 | 3 | 4 | 5 |
| Condition description |  | HEAT | SR: TET (1:1) | SR: TET (2:1) | Neat SR | TET (SR added to cartridge) | HEAT | SR: TET (1:1) | SR: TET (2:1) | Neat SR | TET (SR added to cartridge) |
| Final SR concentration (%) |  | 0% | 12.5% | 16.6% | 25.0% | 50.0% | 0% | 12.5% | 16.6% | 25.0% | 50.0% |
| CFU per 700 µl of sample buffer | 0 | 0/20<sup>#</sup><br>(0) | 0/20<br>(0) | 0/20<br>(0) | 0/19<br>(0) | 0/20<br>(0) | 0/20<br>(0) | 0/20<br>(0) | 0/20<br>(0) | 0/19<br>(0) | 0/20<br>(0) |
|  | 3 | 6/20<br>(30.0) | 6/19<br>(31.6) | 8/18<br>(44.4) | 5/19<br>(26.3) | 5/20<br>(25.0) | 3/20<br>(15.0) | 2/19<br>(10.5) | 5/18<br>(27.8) | 3/19<br>(15.8) | 1/20<br>(5.0) |
|  | 7 | 13/20<br>(65.0) | 13/20<br>(65.0) | 14/18<br>(77.8) | 12/19<br>(63.2) | 7/20<br>(35.0) | 8/20<br>(40.0) | 9/20<br>(45.0) | 10/18<br>(55.6) | 8/19<br>(42.1) | 3/20<br>(15.0) |
|  | 28 | 16/25<br>(64.0) | 24/25<br>(96.0) | 22/25<br>(88.0) | 22/25<br>(88.0) | 10/24<br>(41.7) | 7/25<br>(28.0) | 15/25<br>(60.0) | 16/25<br>(64.0) | 13/25<br>(52.0) | 1/24<br>(4.2) |
|  | 56 | 19/20<br>(95.0) | 20/20<br>(100) | 20/20<br>(100) | 19/19<br>(100) | 20/20<br>(100) | 17/20<br>(85.0) | 20/20<br>(100) | 20/20<br>(100) | 19/19<br>(100) | 17/20<br>(85.0) |
|  | 112 | 18/19<br>(94.7) | - | - | - | 20/20<br>(100) | 18/19<br>(94.74) | - | - | - | 18/20<br>(90.0) |
|  | 336 | 20/20<br>(100) | - | - | - | 19/19<br>(100) | 20/20<br>(100) | - | - | - | 19/19<br>(100) |
|  | Limit of detection (95% CI) | 77.6<br>(51.2-104.0) | 22.7<br>(14.2-31.2) | 30.3<br>(19.9-40.7) | 30.9<br>(21.5-40.3) | 57.1<br>(42.4-71.7) | 95.1<br>(68.9-121.2) | 47.2<br>(35.1-59.2) | 47.0<br>(33.7-60.2) | 51.9<br>(38.5-65.2) | 104.5<br>(80.7-128.3) |
|  | Errors (Total errors/total tests) | 1<sup>a</sup>/145 | 0/104 | 1<sup>b</sup>/102 | 3<sup>b</sup>/104 | 2<sup>c,d</sup>/145 | N/A | N/A | N/A | N/A | N/A |

<sup>#</sup> Number of positive results/total number of samples tested (% positives); Positive test results were defined based on IS6110/IS1081 Ct and *rpoB* Ct for *Mtb* and RIF, respectively. <sup>a</sup>Error 5007: [SPC] probe check failed; <sup>b</sup>Error 2037: The cartridge integrity test failed at valve position
1390; <sup>c</sup>Error 2032: the ultrasonic horn could not be tuned properly; <sup>d</sup>Error 5007: [QC1] probe check failed. CI, confidence interval; CFU, colony forming unit; Ct, cycle threshold; *Mtb*, *Mycobacterium tuberculosis*; N/A, not applicable. RIF, rifampicin; SR, Xpert sample reagent; TET, Tris-EDTA-Tween buffer.

### Performance of Xpert Ultra with sputum matrix versus tongue swabs

Given that tongue swabs processed with heating methods generate elevated pressures within the Xpert cartridge which occasionally cause assays to be aborted as “errors”(1), we undertook an experiment to review the pressure in Xpert cartridges processed with the different swab conditions. However, we did not note errors of this type under any of our test conditions. Sputum samples will often produce high cartridge pressures and error rates when samples are not processed with SR. We therefore sought to assess the Xpert Ultra cartridge pressures obtained on processing sterile swabs dipped in TB-negative sputum with condition 1 and condition 3 protocols at 28 CFU in 700 µl of the relevant buffer.

Swabs containing a sputum matrix and 28 CFU *Mtb* show a trend that was not statistically significant of higher in-cartridge pressures, with median in-cartridge pressures in tests of condition 1 versus condition 3 of 22.2 versus 17.9 psi, p= 0.11. (**Fig. 4a**). The Xpert Ultra tests of sputum matrix swabs detected *Mtb* in 79% of the samples in condition 1 versus 100% detection in condition 3 (p=0.047), and detected RIF susceptibility in 47.4% of sputum matrix swabs in condition 1 versus 85% in condition 3 (p=0.018) (**Table 2**). No false positive results for *Mtb* or RIF susceptibility were observed. Finally, the sputum swab matrix samples had higher mean IS*6110* Ct values in condition 1 versus condition 3 (25.22 versus 22.49, p<0.001). No statistically significant differences were observed between swab matrix or processing method for *rpoB* Ct **(Fig. 4b)** values, likely because many more RIF susceptibility assay results were negative in condition 1 and thus did not produce a Ct value.

**FIG 4.**
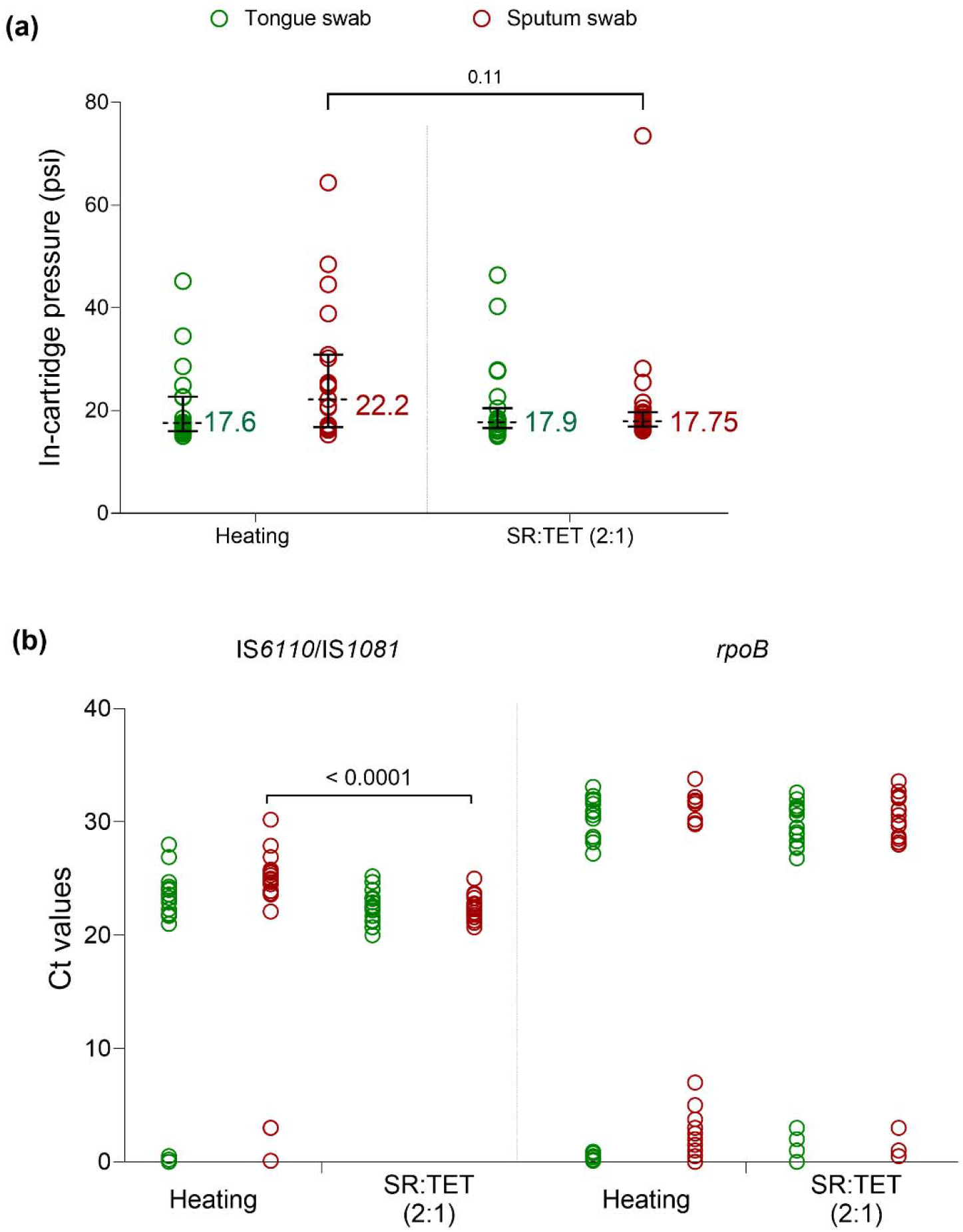
(a) In-cartridge pressure values of tongue and sputum swab samples processed by condition 1 (heat) or condition 3 (diluted SR: TET 2:1); Dotted lines and error bars indicate median and respective 95 % Confidence interval (b) Xpert Ultra Ct values for tongue swab samples processed by condition 1 (heat) or condition 3 (diluted SR: TET 2:1) for *IS*6110/*IS*1081 and *rpoB.* Ct values of 0 were included in the graph to represent the negatives but were not used for statistical analysis. CFU, colony forming unit; Ct, cycle threshold; SR, Xpert sample reagent; TET, Tris-EDTA-Tween buffer. No errors or invalids were observed for this experiment

**TABLE 2.** Xpert Ultra performance with *Mtb* spiked into buffer with swab samples processed using condition 1 (heat) versus condition 3 (diluted SR: TET 2:1) to test impact of sputum matrix and buffer type (TET vs PBS)

| Analyte |  | *Mtb* positivity, n/N (%)<br>(based on IS6110/IS1081 Ct) |  | RIF susceptibility, n/N (%)<br>(based on *rpoB* Ct) |  |
| --- | --- | --- | --- | --- | --- |
| CFU/700 µl | Conditions tested | Condition 1<br>(HEAT) | Condition 3<br>SR: TET (2:1) | Condition 1<br>(HEAT) | Condition 3<br>SR: TET (2:1) |
| **28 CFU** | **Tongue swab** | 16/19 (84.2) | 20/20 (100) | 13/19 (68.4) | 16/20 (80.0) |
|  | **Sputum swab** | 15/19 (79.0) | 20/20 (100) | 9/19 (47.4) | 17/20 (85.0) |
| **28 CFU** | **TET** | N/A | 15/15 (100) | N/A | 13/15 (86.7) |
|  | **PBS** | N/A | 15/15 (100) | N/A | 14/15 (93.3) |

CFU, colony forming unit; Ct, cycle threshold; *Mtb*, *Mycobacterium tuberculosis*; N/A, not applicable. PBS, phosphate buffer saline; RIF, rifampicin; SR, Xpert sample reagent; TET, Tris-EDTA-Tween buffer

### Performance of Xpert Ultra with phosphate buffer saline versus Tris-EDTA-Tween buffer

We compared testing in our Tris-EDTA-Tween buffer versus testing in a PBS buffer, which might be preferred at many test sites, using n=15 samples in each group. All steps for testing with PBS were the same as those for condition 3, except that Tris-EDTA-Tween buffer was replaced with PBS. Xpert Ultra performance was assessed at 28 CFU/700 µl. Detection was not different between the two buffers; Xpert Ultra detected *Mtb* in 100% samples with both PBS and Tris-EDTA-Tween, and detected RIF susceptibility in 93.3% of PBS and 86.7% of Tris-EDTA-Tween samples (p= 1) (**Table 2 and Fig. 5**).

**FIG 5.**
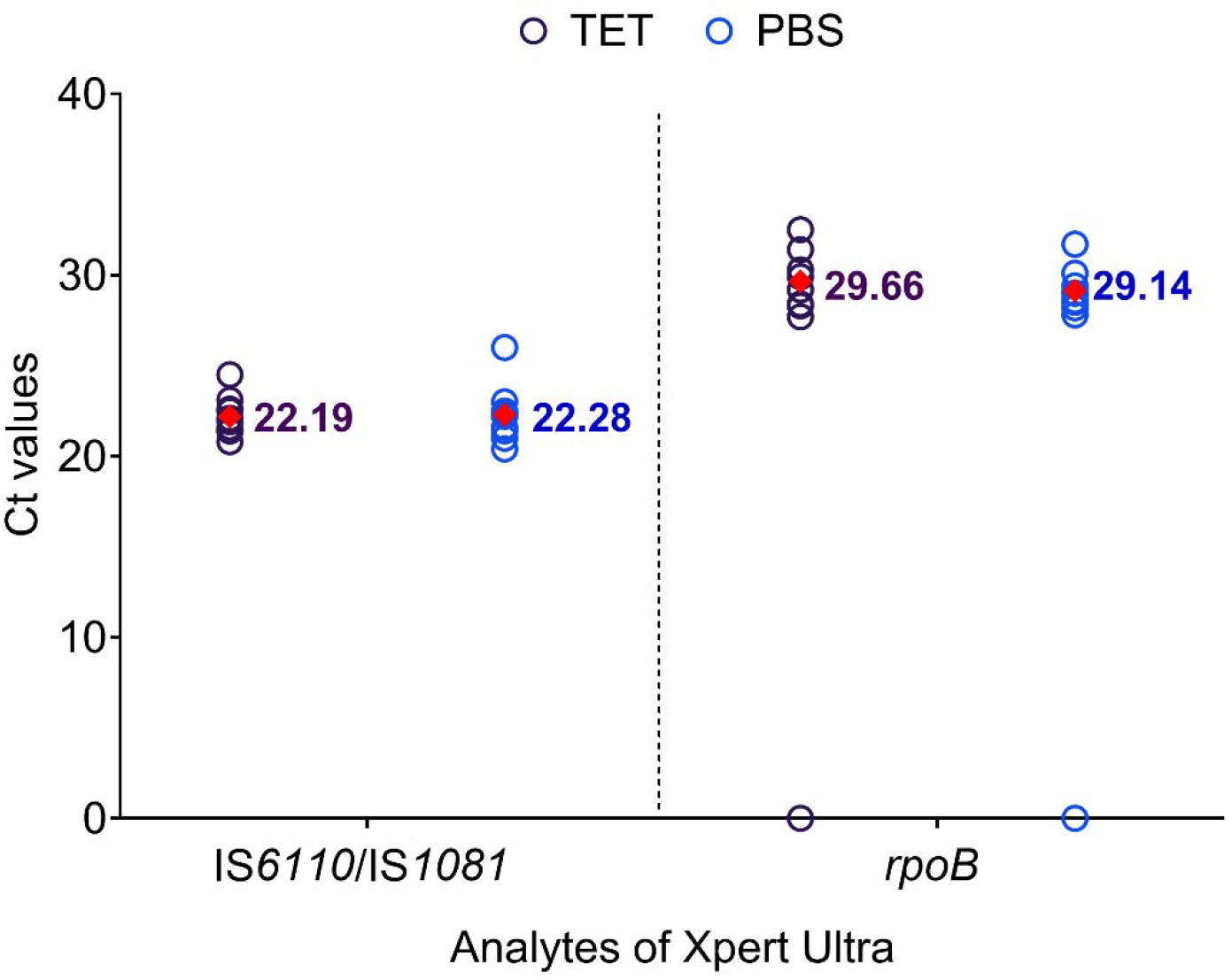
Comparison of IS*6110*/IS*1081* and *rpoB* Ct values at 28 CFU per 700 µl for tongue swab samples processed by condition 3 (diluted SR: TET 2:1) with TET vs PBS as diluent buffer. Mean Ct values are indicated by filled diamonds. Ct values of 0 were included in the graph to represent the negatives but were not used for statistical analysis. Ct, cycle threshold; PBS, phosphate saline buffer; TET, Tris-EDTA-Tween buffer. No errors or invalids were observed for this experiment.

## Discussion

This study demonstrates the utility of several sample processing conditions using diluted Xpert SR to improve the sensitivity of Xpert Ultra to detect TB and RIF resistance in testing tongue swabs of people with presumed TB, compared to the commonly used heat-based protocol. The 2:1 diluted SR buffer (condition 3) had a two-fold lower (i.e. improved) LOD compared to the commonly used heat-based inactivation method and for biosafety reasons was selected for further analysis. There was minimal influence of sputum matrix or dilution buffer on the performance of Xpert Ultra assay using this condition.

In a recent study, processing of tongue swabs with SR before Xpert Ultra testing has resulted in a sensitivity of 78% and specificity of 100%, with sputum as the reference standard (1). The LOD of Xpert Ultra observed in the analytical component of this study was 101.7 and 76.5 CFU/swab with single and double swab SR methods, respectively, wherein the final concentration of SR in the Xpert Ultra cartridge was approximately 66.6% SR (800 µl Tris-EDTA + 1600 µl SR) (1). The sample processing conditions in our study had a final SR concentration ranging from 12.5% to 50%, with 16.6% in our preferred condition 3 resulting in an LOD of 30.3 CFU per 700 µl sample, showing that LOD can be improved using a lower concentration of SR. We chose condition 3 as our preferred condition for subsequent studies of sputum swabs and PBS buffer even though condition 2, which contained 12.5% SR, had a lower LOD of 22.7. We made this selection because the two conditions had virtually identical numerical LODs for RIF susceptibility, and the higher concentration of SR has known sterilizing activity against *Mtb* in sputum (20).

Our study also helped address concerns noted by previous studies on the role of tongue swab components in over-pressurization errors observed with Xpert Ultra testing using the boil inactivation method (19). We confirmed that in-cartridge pressure variations observed with heat-based processing in sputum matrix swabs could be minimized by treatment with diluted SR buffer. Our method may therefore enable improved sensitivity with reduced risk of over-pressurization errors.

Previous oral swab evaluations have used different types of sterile buffer matrices ranging from Tris-EDTA buffer(1, 22), saline(11, 23), sterile lysis buffer (50 mM Tris pH 8.0, 50 mM EDTA, 50 mM sucrose, 100 mM NaCl and 1% SDS) (12, 24, 25), or 7H9+OADC+Tween-80 (26) for storage or dry swab sample processing (1, 2, 11). However, most analytical laboratory evaluations of Xpert Ultra or Xpert MTB/XDR have been performed with TET buffer as a standard test solution for sample processing (27, 28). Hence, we used this buffer to evaluate the LOD of tongue swabs with Xpert Ultra using the modified processing conditions in our study. However, many clinical laboratories in low-resource settings may have difficulty in procuring or making sterile TET buffer. We therefore evaluated the sensitivity of tongue swabs processed with the more readily available PBS. No difference in *Mtb* positivity or RIF susceptibility detection between condition 3 using TET buffer and PBS were observed. As our method also eliminates the need for additional laboratory equipment, the PBS-based condition may be of particular value in low-resource settings. It is likely that PB buffer without saline, as provided with sputum decontamination kits, would work just as well, and assessment of this in clinical specimens is underway.

The Cepheid proprietary sodium hydroxide and isopropanol containing SR was primarily designed to liquify and inactivate sputum samples. SR-inactivated *Mtb* cells are captured on a membrane filter in the Cepheid cartridge during automated sample processing in the Xpert cartridge (27). SR is known to be tuberculocidal (27, 29). Use of an SR-based sample processing method over the existing heat-based protocol therefore has potential to reduce biohazard risk during tongue swab Xpert Ultra processing in a healthcare laboratory setting.

Although we have demonstrated a relatively simple procedure for tongue swab processing for Xpert Ultra testing, this study is limited by the lack of clinical validation of our protocol. It will be essential to study the utility of our processing methodology in tongue swabs collected from people with confirmed TB, smear-negative TB cases based on disease severity, and people with presumptive TB unable to produce sputum. Evaluation of sensitivity and specificity compared with other non-sputum sample types will also be important. However, the strength of our study is in its ability to analytically validate multiple sample processing methods in a systematic manner, which is very difficult to demonstrate in clinical studies.

In conclusion, the performance of Xpert Ultra for detection of *Mtb* positivity and RIF resistance in tongue swabs was improved using a brief incubation in an SR containing buffer, followed by a 3:1 dilution of a non-SR containing buffer when the treated sample is added into the Xpert cartridge, when compared with the existing heat-based protocol. This approach may increase the feasibility of using tongue swabs to screen for TB, thereby expanding the availability of TB testing to additional lower levels of the healthcare system and populations. Further studies are needed to assess the performance of this new tongue swab sample processing condition in a clinical setting.

## Data Availability

All data produced in the present work are contained in the manuscript.

## Acknowledgments

The authors thank Cepheid, Inc. for providing research-use-only Xpert Ultra cartridges and Tris-EDTA-Tween buffer for LOD experiments; however, Cepheid had no role in designing the study or interpreting the results and did not review an advance copy of the manuscript. We also thank the study participants for their time and tongue swab donations. We thank Dr. Paulami Rudra from Rutgers University for her assistance in preparation of reagents for the experiment. Editing support, under the direction of the authors, was provided by Rachel Wright, PhD, funded by Rutgers University, according to Good Publication Practice guidelines.

## References

1. Andama A, Whitman GR, Crowder R, Reza TF, Jaganath D, Mulondo J, Nalugwa TK, Semitala FC, Worodria W, Cook C, Wood RC, Weigel KM, Olson AM, Lohmiller Shaw J, Kato-Maeda M, Denkinger CM, Nahid P, Cangelosi GA, Cattamanchi A. 2022. Accuracy of Tongue Swab Testing Using Xpert MTB-RIF Ultra for Tuberculosis Diagnosis. J Clin Microbiol 60:e0042122.

2. Church EC, Steingart KR, Cangelosi GA, Ruhwald M, Kohli M, Shapiro AE. 2024. Oral swabs with a rapid molecular diagnostic test for pulmonary tuberculosis in adults and children: a systematic review. Lancet Glob Health 12:e45–e54.

3. Wood RC, Luabeya AK, Dragovich RB, Olson AM, Lochner KA, Weigel KM, Codsi R, Mulenga H, de Vos M, Kohli M, Penn-Nicholson A, Hatherill M, Cangelosi GA. 2023. Tongue swab testing on two automated tuberculosis diagnostic platforms, Cepheid Xpert(®) MTB/RIF Ultra and Molbio Truenat(®) MTB Ultima. medRxiv doi:10.1101/2023.10.10.23296833.

4. World Health Organization. Global Tuberculosis Report 2023. https://www.who.int/teams/global-tuberculosis-programme/tb-reports/global-tuberculosis-report-2023. Accessed May 2024.

5. World Health Organization. WHO consolidated guidelines on tuberculosis. Module 3: diagnosis – rapid diagnostics for tuberculosis detection, third edition. https://www.who.int/publications/i/item/9789240089488. Accessed May 2024.

6. Valinetz ED, Cangelosi GA. 2021. A Look Inside: Oral Sampling for Detection of Non-oral Infectious Diseases. J Clin Microbiol 59:e0236020.

7. Swaminathan S, Rekha B. 2010. Pediatric tuberculosis: global overview and challenges. Clin Infect Dis 50 Suppl 3:S184–94.

8. Mendelson M. 2007. Diagnosing tuberculosis in HIV-infected patients: challenges and future prospects. Br Med Bull 81–82:149-165.

9. Carbone AdSS, Paião DSG, Sgarbi RVE, Lemos EF, Cazanti RF, Ota MM, Junior AL, Bampi JVB, Elias VPF, Simionatto S, Motta-Castro ARC, Pompílio MA, de Oliveira SMdV, Ko AI, Andrews JR, Croda J. 2015. Active and latent tuberculosis in Brazilian correctional facilities: a cross-sectional study. BMC Infect Dis 15:24.

10. Venturini E, Turkova A, Chiappini E, Galli L, de Martino M, Thorne C. 2014. Tuberculosis and HIV co-infection in children. BMC Infect Dis 14 Suppl 1:S5.

11. Cox H, Workman L, Bateman L, Franckling-Smith Z, Prins M, Luiz J, Van Heerden J, Ah Tow Edries L, Africa S, Allen V, Baard C, Zemanay W, Nicol MP, Zar HJ. 2022. Oral Swab Specimens Tested With Xpert MTB/RIF Ultra Assay for Diagnosis of Pulmonary Tuberculosis in Children: A Diagnostic Accuracy Study. Clin Infect Dis 75:2145-2152.

12. Lima F, Santos AS, Oliveira RD, Silva CCR, Gonçalves CCM, Andrews JR, Croda J. 2020. Oral swab testing by Xpert® MTB/RIF Ultra for mass tuberculosis screening in prisons. J Clin Tuberc Other Mycobact Dis 19:100148.

13. Mesman AW, Calderon R, Soto M, Coit J, Aliaga J, Mendoza M, Franke MF. 2019. Mycobacterium tuberculosis detection from oral swabs with Xpert MTB/RIF ULTRA: a pilot study. BMC Research Notes 12:349.

14. Steadman A, Andama A, Ball A, Mukwatamundu J, Khimani K, Mochizuki T, Asege L, Bukirwa A, Kato JB, Katumba D, Kisakye E, Mangeni W, Mwebe S, Nakaye M, Nasuna I, Nyawere J, Visente D, Cook C, Nalugwa T, Bachman CM, Semitalia F, Weigl BH, Connelly J, Worodria W, Cattamanchi A. 2023. New manual qPCR assay validated on tongue swabs collected and processed in Uganda shows sensitivity that rivals sputum-based molecular TB diagnostics. medRxiv doi:10.1101/2023.08.10.23293680.

15. Bahr NC, Nuwagira E, Evans EE, Cresswell FV, Bystrom PV, Byamukama A, Bridge SC, Bangdiwala AS, Meya DB, Denkinger CM, Muzoora C, Boulware DR. 2018. Diagnostic accuracy of Xpert MTB/RIF Ultra for tuberculous meningitis in HIV-infected adults: a prospective cohort study. Lancet Infect Dis 18:68–75.

16. Mansfield M, McLaughlin AM, Roycroft E, Montgomery L, Keane J, Fitzgibbon MM, Rogers TR. 2022. Diagnostic Performance of Xpert MTB/RIF Ultra Compared with Predecessor Test, Xpert MTB/RIF, in a Low TB Incidence Setting: a Retrospective Service Evaluation. Microbiol Spectr 10:e0234521.

17. Chakravorty S, Simmons AM, Rowneki M, Parmar H, Cao Y, Ryan J, Banada PP, Deshpande S, Shenai S, Gall A, Glass J, Krieswirth B, Schumacher SG, Nabeta P, Tukvadze N, Rodrigues C, Skrahina A, Tagliani E, Cirillo DM, Davidow A, Denkinger CM, Persing D, Kwiatkowski R, Jones M, Alland D. 2017. The New Xpert MTB/RIF Ultra: Improving Detection of *Mycobacterium tuberculosis* and Resistance to Rifampin in an Assay Suitable for Point-of-Care Testing. mBio 8:10.1128/mbio.00812-17.

18. Hoel IM, Syre H, Skarstein I, Mustafa T. 2020. Xpert MTB/RIF ultra for rapid diagnosis of extrapulmonary tuberculosis in a high-income low-tuberculosis prevalence setting. Sci Rep 10:13959.

19. Whitman GR. 2021. Tongue dorsum swab processing for the detection of tuberculosis with Cepheid Xpert® MTB/RIF Ultra. University of Washington.

20. Helb D, Jones M, Story E, Boehme C, Wallace E, Ho K, Kop J, Owens MR, Rodgers R, Banada P, Safi H, Blakemore R, Lan NT, Jones-Lopez EC, Levi M, Burday M, Ayakaka I, Mugerwa RD, McMillan B, Winn-Deen E, Christel L, Dailey P, Perkins MD, Persing DH, Alland D. 2010. Rapid detection of Mycobacterium tuberculosis and rifampin resistance by use of on-demand, near-patient technology. J Clin Microbiol 48:229–37.

21. Cox H, Workman L, Bateman L, Franckling-Smith Z, Prins M, Luiz J, Van Heerden J, Ah Tow Edries L, Africa S, Allen V, Baard C, Zemanay W, Nicol MP, Zar HJ. 2022. Oral Swab Specimens Tested With Xpert MTB/RIF Ultra Assay for Diagnosis of Pulmonary Tuberculosis in Children: A Diagnostic Accuracy Study. Clinical Infectious Diseases 75:2145-2152.

22. Shapiro AE, Olson AM, Kidoguchi L, Niu X, Ngcobo Z, Magcaba ZP, Ngwane MW, Whitman GR, Weigel KM, Wood RC, Wilson DPK, Drain PK, Cangelosi GA. 2022. Complementary Nonsputum Diagnostic Testing for Tuberculosis in People with HIV Using Oral Swab PCR and Urine Lipoarabinomannan Detection. Journal of Clinical Microbiology 60:e00431–22.

23. Song Y, Ma Y, Liu R, Shang Y, Ma L, Huo F, Li Y, Shu W, Wang Y, Gao M, Pang Y. 2021. Diagnostic Yield of Oral Swab Testing by TB-LAMP for Diagnosis of Pulmonary Tuberculosis. Infection and Drug Resistance 14:89–95.

24. Wood RC, Andama A, Hermansky G, Burkot S, Asege L, Job M, Katumba D, Nakaye M, Mwebe SZ, Mulondo J, Bachman CM, Nichols KP, Le Ny A-LM, Ortega C, Olson RN, Weigel KM, Olson AM, Madan D, Bell D, Cattamanchi A, Worodria W, Semitala FC, Somoskovi A, Cangelosi GA, Minch KJ. 2021. Characterization of oral swab samples for diagnosis of pulmonary tuberculosis. PLOS ONE 16:e0251422.

25. Wood RC, Luabeya AK, Weigel KM, Wilbur AK, Jones-Engel L, Hatherill M, Cangelosi GA. 2015. Detection of Mycobacterium tuberculosis DNA on the oral mucosa of tuberculosis patients. Scientific Reports 5:8668.

26. Ealand C, Peters J, Jacobs O, Sewcharran A, Ghoor A, Golub J, Brahmbhatt H, Martinson N, Dangor Z, Lala SG, Kana B. 2021. Detection of Mycobacterium tuberculosis Complex Bacilli and Nucleic Acids From Tongue Swabs in Young, Hospitalized Children. Frontiers in Cellular and Infection Microbiology 11.

27. Helb D, Jones M, Story E, Boehme C, Wallace E, Ho K, Kop J, Owens MR, Rodgers R, Banada P, Safi H, Blakemore R, Lan NTN, Jones-López EC, Levi M, Burday M, Ayakaka I, Mugerwa RD, McMillan B, Winn-Deen E, Christel L, Dailey P, Perkins MD, Persing DH, Alland D. 2010. Rapid Detection of *Mycobacterium tuberculosis* and Rifampin Resistance by Use of On-Demand, Near-Patient Technology. J Clin Microbiol 48:229–237.

28. Cao Y, Parmar H, Gaur RL, Lieu D, Raghunath S, Via N, Battaglia S, Cirillo DM, Denkinger C, Georghiou S, Kwiatkowski R, Persing D, Alland D, Chakravorty S. 2021. Xpert MTB/XDR: a 10-Color Reflex Assay Suitable for Point-of-Care Settings To Detect Isoniazid, Fluoroquinolone, and Second-Line-Injectable-Drug Resistance Directly from Mycobacterium tuberculosis-Positive Sputum. J Clin Microbiol 59:10.1128/jcm.02314-20.

29. Banada PP, Sivasubramani SK, Blakemore R, Boehme C, Perkins MD, Fennelly K, Alland D. 2010. Containment of bioaerosol infection risk by the Xpert MTB/RIF assay and its applicability to point-of-care settings. J Clin Microbiol 48:3551–7.

